# Reducing Inequalities Using an Unbiased Machine Learning Approach to Identify Births with the Highest Risk of Preventable Neonatal Deaths

**DOI:** 10.1101/2024.01.12.24301163

**Authors:** Antonio P. Ramos, Fabio Caldieraro, Marcus L. Nascimento, Raphael Saldanha

**Affiliations:** California Population Center, University of California, Los Angeles, USA; Pensi Institute, São Paulo, Brazil; Brazilian School of Public and Business Administration, Getulio Vargas Foundation, Rio de Janeiro, Brazil; School of Applied Mathematics, Getulio Vargas Foundation, Rio de Janeiro, Brazil; Institute of Scientific and Technological Communication and Information in Health, Oswaldo Cruz Foundation, Rio de Janeiro, Brazil

**Keywords:** Algorithmic Bias, Health Inequality, Machine Learning, Neonatal Mortality, Program Targeting

## Abstract

**Background:** Despite contemporaneous declines in neonatal mortality, recent studies show the existence of left-behind populations that continue to have higher mortality rates than the national averages. Additionally, many of these deaths are from preventable causes. This reality creates the need for more precise methods to identify high-risk births, allowing policymakers to target them more effectively. This study fills this gap by developing unbiased machine-learning approaches to more accurately identify births with a high risk of neonatal deaths from preventable causes.

**Methods:** We link administrative databases from the Brazilian health ministry to obtain birth and death records in the country from 2015 to 2017. The final dataset comprises 8,797,968 births, of which 59,615 newborns died before reaching 28 days alive (neonatal deaths). These neonatal deaths are categorized into preventable deaths (42,290) and non-preventable deaths (17,325). Our analysis identifies the death risk of the former group, as they are amenable to policy interventions. We train six machine-learning algorithms, test their performance on unseen data, and evaluate them using a new policy-oriented metric. To avoid biased policy recommendations, we also investigate how our approach impacts disadvantaged populations.

**Results:** XGBoost was the best-performing algorithm for our task, with the 5% of births identified as highest risk by the model accounting for over 85% of the observed deaths. Furthermore, the risk predictions exhibit no statistical differences in the proportion of actual preventable deaths from disadvantaged populations, defined by race, education, marital status, and maternal age. These results are similar for other threshold levels.

**Conclusions:** We show that, by using publicly available administrative data sets and ML methods, it is possible to identify the births with the highest risk of preventable deaths with a high degree of accuracy. This is useful for policymakers as they can target health interventions to those who need them the most and where they can be effective without producing bias against disadvantaged populations. Overall, our approach can guide policymakers in reducing neonatal mortality rates and their health inequalities. Finally, it can be adapted for use in other developing countries.

## 1 Background

In recent years, many countries have achieved considerable progress in reducing early-life mortality (ELM), and many are in line to achieve the United Nations’ Sustainable Development Goals (SDGs). These reductions are important and associated with improved health outcomes^1,2^. However, health disparities remain high, even in countries in line to achieve the SDGs. These disparities may exist between ethnic groups, geographic regions, and levels of education, to mention a few subgroups ^3,4^. This is particularly concerning for deaths from preventable causes, where available interventions could be used^5^.

International agencies and local policymakers have recognized these disparities between subgroups^6,7^. The most common approach to identifying high-risk groups has been stratifying mortality rates by subgroups, such as gender, socioeconomic status, and geographic location. While useful for some purposes, these approaches ignore within-group variability, whereas children from the same subgroup may have very different mortality rates. Recent studies showed that within-group variability is higher than between-group variability ^8,9^.

The decline in mortality rates makes it even more useful to adopt methods that can precisely identify those who still have a high risk of preventable deaths. This is particularly salient when only a fraction of the population can be given the needed intervention because of two factors. First, in many contexts in the developing world, resources are scarce. At the same time, at-risk individuals may demand considerable attention; thus, the importance of not squandering resources on those who do not truly need them. Secondly, the smaller the population that can receive an intervention, the more difficult the task of correctly identifying the individuals to be targeted. In this paper, we develop and explore a new approach. Using a large administrative data set with individual-level information about each birth, we employ machine learning models (ML) to estimate the risk of preventable neonatal death for new unseen births.

Our goal is to develop a data-driven approach that combines several risk factors and provides digested information that identifies which neonates are at the highest risk of preventable death.

In addition, we apply a new metric to evaluate the performance of the machine learning models developed. Our proposed approach does not use traditional criteria such as specificity and accuracy, or F1 metrics that are difficult to interpret. Instead, our metric assesses how effectively our algorithms identify births at high risk of death due to preventable causes.

We also address concerns of bias in ML algorithms, given recent literature that shows the potential risk that the application of these methods can be more favorable to privileged populations^10,11,12,13,14^. Our models do not exhibit this behavior and capture similar proportions of preventable neonatal deaths from advantaged and less-advantaged populations.

## 2 Methods

### 2.1 Approach

In this research, the unit of analysis is the individual birth. We aim to identify the births with the highest neonatal mortality risk from preventable causes. As such, we included all the available information from the administrative databases, which improved the precision of our targeting.

### 2.2 Data sources

We use administrative databases from the Brazilian health ministry to obtain birth and death records in the entire country from 2015 to 2017 and information about health facilities, professionals, and available equipment. All data is available at https://datasus.saude.gov.br. Still, it is organized into three different health information systems: SINASC (Sistema de Informações sobre Nascidos Vivos), SIM (Sistema de Informação sobre Mortalidade), CNES (Cadastro Nacional de Estabelecimentos de Saúde), which we describe below.

SINASC includes all live births in the Brazilian territory, recording epidemiological and administrative information about the mothers and children. SIM, in turn, includes all deaths in the territory, containing epidemiological and administrative information and their circumstances. Fetal deaths are not considered as they are beyond the scope of this paper. Finally, CNES records a snapshot of Brazilian health facilities at a point in time. These systems contain three tables with all live births, deaths, and health facilities information.

To merge SIM and SINASC data, we used the field NUMERODN. It contains a unique number identifying each live birth. Records on SIM contain this information in cases of deaths within the first year since birth. Subsequently, we merged the information with CNES data by the CNES number, a unique identifier for health facilities in both the SINASC and CNES databases. The resulting raw dataset totals 8,829,944 records.

When merging the three databases, we identified and removed duplicate observations in the SIM and SINASC tables to avoid inconsistencies. With the deduplicated tables, deterministic linkages were executed.^1^

We note that we also performed linkages using probabilistic matching according to the method proposed by Enamorado et al. (2019). Although the probabilistic matching enabled us to link a larger proportion of records, we found that the use of probabilistic matching would not improve the predictive power of the algorithm; thus, for the sake of parsimony, we used the dataset without

In the raw dataset, a few additional treatments were performed. SIM records that were not linked to a SINASC record were not considered. Moreover, we did not consider a few residual records with no birthdate and records in which the difference between the birthdate and the date of death was negative. After these treatments, the linkage between SIM and SINASC had a linkage success of 78.78%, whereas the linkage with CNES had a linkage success of 96.83%. The resulting cleaned dataset comprises 8,797,968 births and 59,615 neonatal deaths.

### 2.3 Feature Engineering

Our set of features^16^ consists of the following variables: place of delivery, health facility type, maternal age at birth, sex, 1-min Apgar score, 5-min Apgar score, birth weight, gestational age, week of gestation, pregnancy type, delivery type, maternal education, presence of congenital anomaly, maternal ethnicity, antenatal visits, month of first antenatal visit, presentation type, induced labor, professional that assisted the labor, number of previous live births, number of previous fetal losses and abortions, number of previous pregnancies, number of previous vaginal deliveries, number of previous cesarean deliveries. In addition, we have also used marital status and state of birth (the definition and type of each of these features are in Table 1)

**Table 1:**
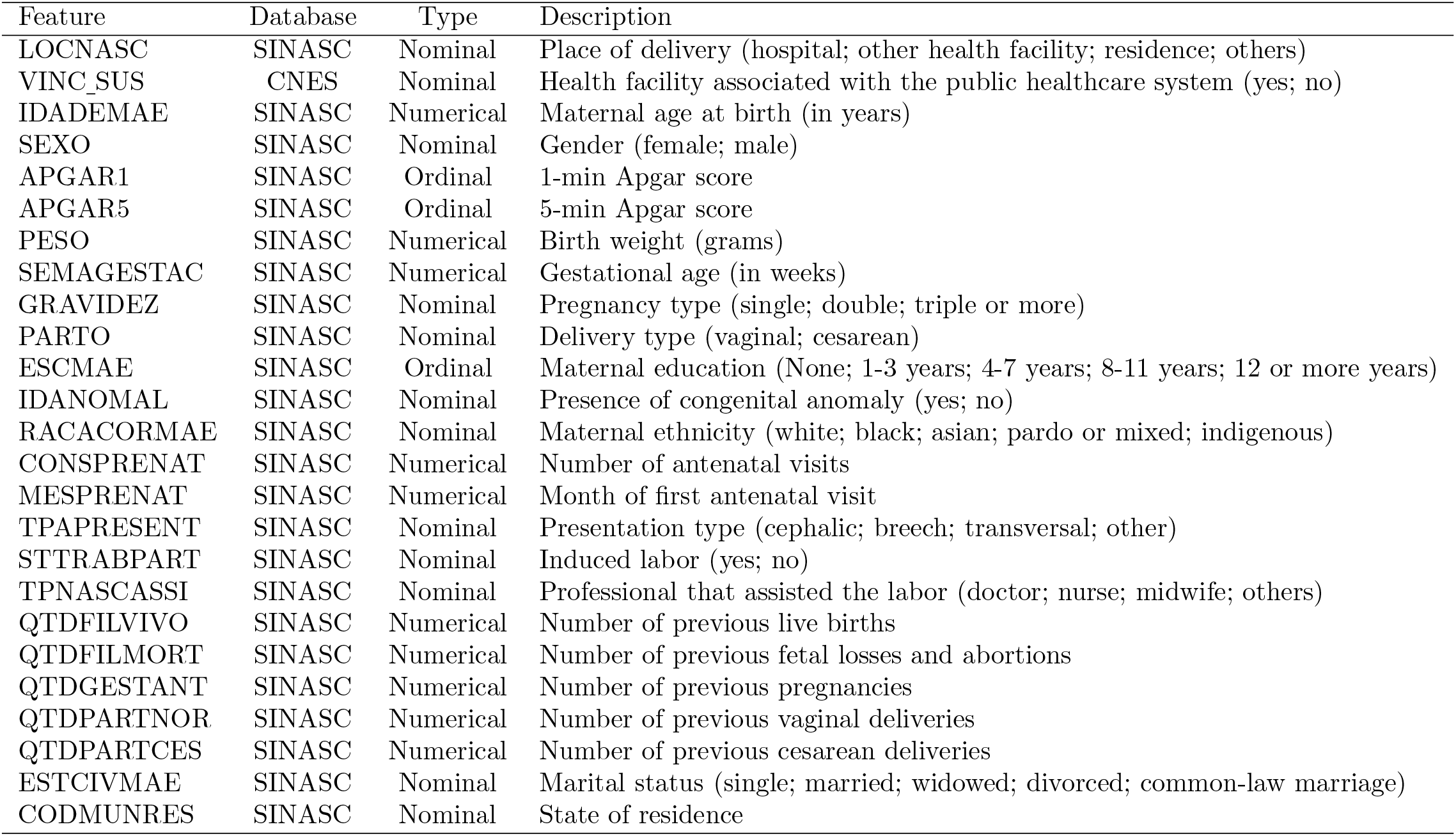
Features information: administrative database, type, and description.

We analyze a nominal categorical target variable with three possible outcomes: alive, preventable death^17^, and non-preventable death. Among the non-preventable deaths, we have external causes of death and ill-defined deaths. The number of preventable deaths is 42,290, whereas the number of non-preventable deaths is 17,325.

To improve analysis efficiency, categorical variables were stored with codes. We did so by performing a relabeling procedure guided by the data dictionaries issued by the DataSUS, using the package *microdatasus* ^18^. We treated missing data via imputation and applied the package *Amelia* ^19^. Both packages are available in the R Statistical Software repository^20^.

As a pre-processing procedure, we centered and scaled the data, by subtracting the mean and dividing by the standard deviation. We also identified and excluded features with zero or near zero variance. Finally, we filtered out highly correlated features. Details are available upon request.

### 2.4 Modeling

The final dataset is partitioned into training and test sets: 7,038,375 observations (80.00% of the total) are used to train six different machine learning algorithms, while 1,759,593 observations (20.00% of the total) are used to evaluate the performance of our targeting criterion on new unseen data.

We estimated preventable neonatal mortality risk for each birth in the data set through flexible ML methods that use the above features. These methods were logistic regression, least absolute shrinkage and selection operator regression (LASSO), elastic-net regularized logistic regression (elastic net), random forest (RF), extreme gradient boosting over trees (XGBoost), and neural networks (NNs). We used the package *caret* available in the R Statistical Software^20^ to run the machine learning algorithms.

Logistic regression^21^ is the standard estimation of a linear model that estimates the parameters *β*_*j*_ for each feature *j* to maximize a logistic likelihood function by minimizing the negative log-likelihood. LASSO^22^ is essentially an implementation of linear regression that uses a 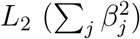 norm penalty to regularize or “shrink” the model, preventing overfitting. It is similar to the logistic regression but includes a penalty term equal to 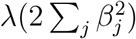, where the parameter *λ* is a non-negative real number that determines the strength of the regulation.

Elastic net^23^ combines *L*_1_ norm (∑_*j*_ |*β*_*j*_|) and 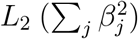 norm penalties to regularize the model. It minimizes the negative log-likelihood plus a penalty term equals to 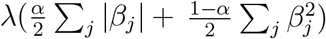, where the parameters *α* and *λ* are defined on the unit interval and on the non-negative real numbers respectively. As particular cases, elastic net comprises LASSO regression (*α* = 1) and logistic regression (*λ* = 0).

Our application first tested a cross-validation procedure to choose the parameters *α* and *λ* in elastic net and the parameter *λ* in LASSO. However, their performances were not close to the logistic regression. For that purpose, we fixed *α* = 0.5 and *λ* = 0.001 in elastic net, and *λ* = 0.001 in LASSO. The method *glmnet* was used for all three algorithms.

The methods RF^24^ and XGBoost^25^ are tree-based algorithms. The simplest tree-based algorithms are classification and regression trees (CART^26^). Both single-tree models recursively group the outcome observations with similar values using cutoff values of the features. Although single-tree models are easy to interpret, their performance is frequently poor and very sensitive to small changes in the input data. By combining several trees, RF and XGBoost methods improve single-tree algorithm performance. The former averages the estimates of a set of trees, each obtained from a random subset of features and trained on a random subset of the observations. The latter also combines several trees, but it initiates with one tree, and new trees are iteratively trained on the errors of the prior set of trees.

As applied by us, in RF, the *ranger* method was employed, and (i) each forest encompasses 500 trees, (ii) the number of variables randomly sampled for each tree split (`mtry`) was set to 5 (the square root of the number of features), (iii) the minimal node size (`min.node.size`) was set to 1, and (iv) we choose the gini index as splitting rule (`splitrule`). In XGBoost, the *xgbTree* method was employed, and (i) the number of iterations for the boosting procedure (`nrounds`) was set to 250, (ii) the learning rate (*η* ∈ (0, 1) was set to 0.3 to prevent over-fitting, (iii) the maximum depth of the trees (`max_depth`) was set to 4, (iv) the proportion of the variables to be considered for tree construction (`colsample_bytree`) was set to the interval (0.6, 1), and the proportion of observations from the training set used for modeling (`subsample`) was set to the interval (0.5, 1).

The NN methods^27^ are constituted by an output layer and node layers, including an input layer and one or more hidden layers. The input layer takes the features, and no processing is done. All kinds of processing are executed on the hidden layers and transferred to the output layer. The output layer, in turn, is the final layer, bringing the final value resulting from the learning process in the hidden layers. The nodes, also known as artificial neurons, are connected, and these associations are characterized by their weights, thresholds, and activation functions. Nodes are activated, and data are sent to the next network layer when their outputs exceed a specified threshold value. Otherwise, no data is transmitted to the next layer.

Although we tested specifications with more than one hidden layer using the *mlpML* method, they performed similarly to the neural network with only one hidden layer. Thus, our application employed the *mlp* method, specifying a layer with 25 nodes.

### 2.5 Performance Metrics

For our task, we did not find it useful to adopt traditional prediction performance metrics, such as classification accuracy, confusion matrices, specificity/sensitivity statistics, or precision/recall statistics, all of which require a threshold for deciding when a risk score is high enough to merit a warning. These can be misleading when applied to rare outcomes, as in the problem we focus on. In our case, if we predicted no neonatal mortality, that model would be right 99% of the time, yet it would be useless, as it wouldn’t allow us to identify those who could be targeted. We neither find it useful to adopt “threshold-free” approaches that report accuracy in a way that does not depend on choosing one threshold, such as ROC-AUC and F-scores do, because they are difficult to give any valuable policy meaning in our context.

We instead recognize that if one has a resource constraint—only a certain fraction of cases one can act on—it gives a reason to compute the proportion of deaths captured by setting the threshold levels of the highest predicted mortality risk. For example, suppose we imagine that a policymaker can only provide intervention to the 5% (or 10%) who need it the most. In this case, the threshold can be set to whatever fraction of high-risk births they have resources for targeting. An appropriate approach, therefore, can concentrate a substantial amount of neonatal deaths in small percentages of high-risk individuals.

### 2.6 Algorithmic Bias

Algorithmic bias ^10,11,12,13,14^ is a well-documented problem with striking implications for health care and public policy. Therefore, besides concentrating a substantial amount of neonatal preventable deaths in small percentages of high-risk individuals, our targeting criterion should also be able to avoid disadvantaging the most vulnerable groups.

To check whether our preferred model would not disadvantage the most vulnerable populations, we checked its performance for four different sub-groups identified using the demographic variables in our dataset. These sub-groups are newborns from *non-white mothers, low-education mothers, underage mothers*, and *single mothers*. We used the test sample as a reference and compared its composition with individuals with the highest predicted risk of neonatal preventable death for different threshold levels. For that, we construct confidence intervals based on the Normal approximation for the mortality rate of each group and check whether these intervals contain their respective mortality rates in the test sample. We also perform hypothesis tests to verify whether the proportions of preventable deaths captured by the algorithm 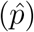 are statistically equal to the proportion of preventable deaths in the test sample (*p*_0_). The null hypothesis is 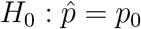 and the alternative hypothesis, 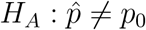.

## 3 Results

Recall that we calculate our performance metric by setting the highest predicted mortality risk threshold levels and considering the percentage of neonatal preventable deaths in the test sample for each threshold level. Figure 1 summarizes the results.

**Figure 1:**
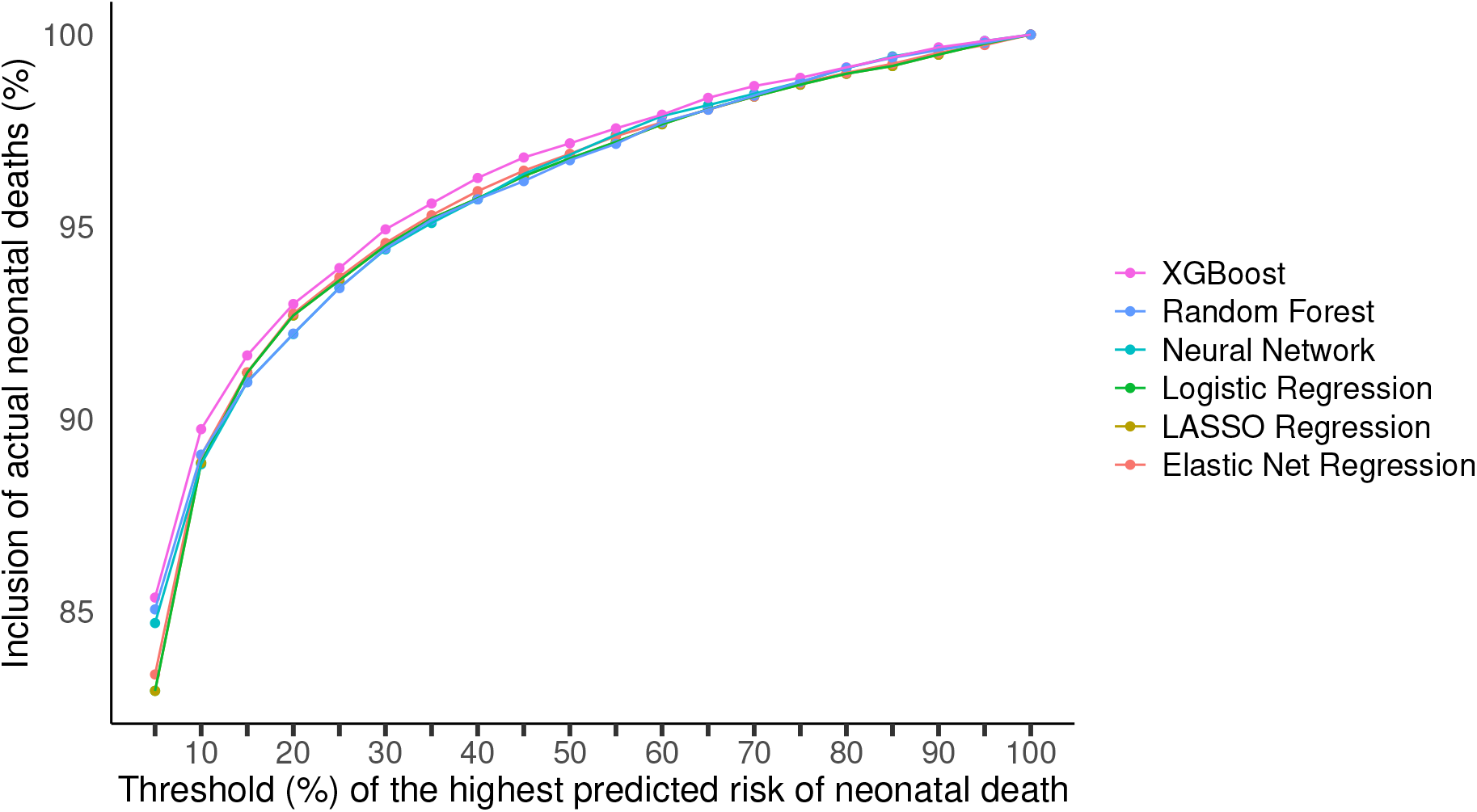
Performance of the different ML methods.

Our best model in terms of predictive performance is the XGBoost method. With that algorithm, in our test sample, including the 5% highest risk births, our model captures 85% of preventable neonatal deaths. The XGBoost is never worse than other competing methods and thus is selected as our preferred model. Traditional performance metrics are reported in Table 2.

**Table 2:** Predictive performance for preventable neonatal mortality on the test set for each machine learning algorithm.

| Performance measure | Machine learning algorithm |  |  |  |  |  |
| --- | --- | --- | --- | --- | --- | --- |
|  | Neural Network | Extreme Gradient Boosting | LASSO Regression | Elastic Net Regression | Logistic Regression | Random Forest |
| Balanced Accuracy | 0.8450 | 0.8353 | 0.8268 | 0.8302 | 0.8268 | 0.8361 |
| Sensitivity | 0.7058 | 0.6840 | 0.6658 | 0.6729 | 0.6658 | 0.6850 |
| Specificity | 0.9842 | 0.9866 | 0.9879 | 0.9875 | 0.9879 | 0.9871 |
| PPV | 0.1780 | 0.1988 | 0.2100 | 0.2068 | 0.2100 | 0.2051 |
| NPV | 0.9985 | 0.9984 | 0.9983 | 0.9984 | 0.9983 | 0.9984 |
| Recall | 0.7058 | 0.6840 | 0.6658 | 0.6729 | 0.6658 | 0.6850 |
| F1-Score | 0.2843 | 0.3081 | 0.3193 | 0.3164 | 0.3193 | 0.3157 |

As a robustness check, we tested the performance of this model to capture “future” births, we retrained the model using the early 80% of the births (using data from January 2015 to May 2017) and tested the model in the later 20% of the births (using data from June 2017 to December 2017). The results show that the model performs nominally better in predicting later births than random births, although the differences in performance are not statistically significant. These results are presented in Figure 2. Comparative performance based on our metric is in Table 3, whereas traditional performance metrics are in Table 4.

**Table 3:** Performance of the XGBoost algorithm considering random and time-based splits and their corresponding 95% uncertainty intervals (UI).

| Threshold<br>(%) | Random split |  | Time-based split |  |
| --- | --- | --- | --- | --- |
|  | Neonatal deaths (%) | UI | Neonatal deaths (%) | UI |
| 5 | 85.37 | (84.65,86.10) | 86.29 | (85.49,87.09) |
| 10 | 89.75 | (89.12,90.38) | 90.34 | (89.67,91.02) |
| 15 | 91.66 | (91.09,92.24) | 92.36 | (91.75,92.97) |
| 20 | 93.00 | (92.47,93.53) | 93.57 | (93.01,94.14) |
| 25 | 93.93 | (93.43,94.44) | 94.38 | (93.86,94.90) |
| 30 | 94.94 | (94.48,95.40) | 95.19 | (94.71,95.67) |
| 35 | 95.61 | (95.18,96.04) | 95.88 | (95.45,96.32) |
| 40 | 96.28 | (95.88,96.67) | 96.44 | (96.04,96.84) |
| 45 | 96.81 | (96.43,97.19) | 96.88 | (96.50,97.26) |
| 50 | 97.17 | (96.82,97.53) | 97.34 | (96.99,97.69) |
| 55 | 97.56 | (97.24,97.89) | 97.71 | (97.38,98.05) |
| 60 | 97.92 | (97.62,98.22) | 98.04 | (97.73,98.35) |
| 65 | 98.36 | (98.09,98.63) | 98.30 | (98.01,98.58) |
| 70 | 98.66 | (98.42,98.91) | 98.65 | (98.39,98.91) |
| 75 | 98.88 | (98.65,99.11) | 99.08 | (98.87,99.29) |
| 80 | 99.15 | (98.94,99.35) | 99.29 | (99.11,99.48) |
| 85 | 99.41 | (99.24,99.58) | 99.52 | (99.37,99.67) |
| 90 | 99.67 | (99.54,99.80) | 99.68 | (99.56,99.81) |
| 95 | 99.83 | (99.75,99.92) | 99.85 | (99.76,99.93) |

**Table 4:** Predictive performance for preventable neonatal mortality on the test set for the XGBoost algorithm, considering random and time-based splits.

| Performance measure | Extreme Gradient Boosting (XGBoost) |  |
| --- | --- | --- |
|  | Random split | Time-based split |
| Balanced Accuracy | 0.8353 | 0.8440 |
| Sensitivity | 0.6840 | 0.7010 |
| Specificity | 0.9866 | 0.9869 |
| PPV | 0.1988 | 0.2025 |
| NPV | 0.9984 | 0.9985 |
| Recall | 0.6840 | 0.7010 |
| F1-Score | 0.3081 | 0.3143 |

**Figure 2:**
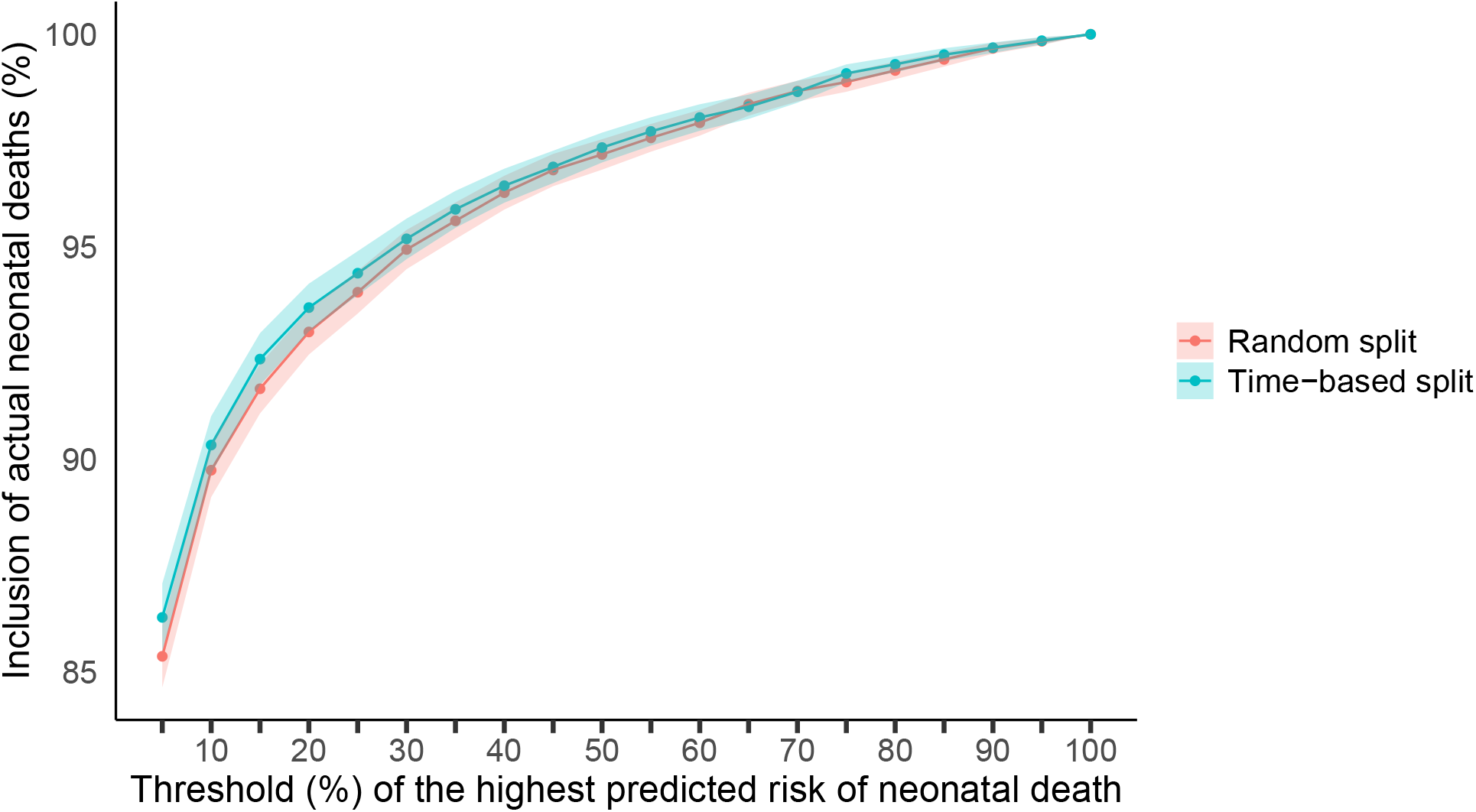
Performance of the XGBoost algorithm considering random and time-based splits and their corresponding 95% uncertainty intervals.

To explore how valuable each variable is in influencing the performance of our predictive model, we performed a Shapley additive explanations (SHAP) analysis ^28^. The SHAP analysis reveals that the factors that influence the risk predictions are the Birth weight, Number of antenatal visits, Gestational age, Number of previous live births, and the Apgar scores. These results are depicted in Figure 5.

One of the main objectives of our model is to propose a method that is not biased against vulnerable populations. Thus, we checked the model performance for the four different subgroups of newborns from disadvantaged populations as presented in the *Algorithmic Bias* Subsection.

In the first analysis, we compared the percentage of disadvantaged individuals selected as high-risk versus the proportion of disadvantaged individuals in our test sample for each sub-group. Table 5 reports these results, which are also depicted in Figure 3.

**Table 5:** Proportions of individuals selected by the algorithm.

| Threshold<br>(%) | n | Maternal ethnicity<br>($p_0 = 63.90\%$) | | | Maternal education<br>($p_0 = 20.22\%$) | | | Age of the mother<br>($p_0 = 5.00\%$) | | | Marital status<br>($p_0 = 44.04\%$) | | |
| --- | --- | --- | --- | --- | --- | --- | --- | --- | --- | --- | --- | --- | --- |
| | | $\hat{p}$ (%) | 95% CI | | $\hat{p}$ (%) | 95% CI | | $\hat{p}$ (%) | 95% CI | | $\hat{p}$ (%) | 95% CI | |
| 5 | 87,979 | 70.56 | (70.26,70.86) |  | 26.70 | (26.41,27.00) |  | 7.59 | (7.41,7.76) |  | 49.24 | (48.91,49.57) |  |
| 10 | 175,959 | 72.75 | (72.54,72.96) |  | 29.61 | (29.40,29.82) |  | 7.52 | (7.40,7.65) |  | 48.84 | (48.61,49.08) |  |
| 15 | 263,938 | 73.82 | (73.65,73.99) |  | 30.52 | (30.34,30.69) |  | 7.54 | (7.44,7.64) |  | 48.76 | (48.57,48.95) |  |
| 20 | 351,918 | 74.38 | (74.24,74.53) |  | 31.06 | (30.91,31.22) |  | 7.49 | (7.41,7.58) |  | 48.68 | (48.51,48.84) |  |
| 25 | 439,898 | 74.69 | (74.56,74.82) |  | 31.14 | (31.01,31.28) |  | 7.42 | (7.34,7.50) |  | 48.64 | (48.50,48.79) |  |
| 30 | 527,877 | 74.79 | (74.68,74.91) |  | 30.98 | (30.86,31.11) |  | 7.35 | (7.28,7.42) |  | 48.56 | (48.43,48.70) |  |
| 35 | 615,857 | 74.72 | (74.61,74.83) |  | 30.65 | (30.53,30.76) |  | 7.25 | (7.18,7.31) |  | 48.47 | (48.34,48.59) |  |
| 40 | 703,837 | 74.54 | (74.44,74.65) |  | 30.20 | (30.09,30.31) |  | 7.15 | (7.09,7.21) |  | 48.34 | (48.22,48.45) |  |
| 45 | 791,816 | 74.20 | (74.10,74.30) |  | 29.65 | (29.55,29.75) |  | 7.04 | (6.98,7.10) |  | 48.20 | (48.09,48.31) |  |
| 50 | 879,796 | 73.72 | (73.63,73.81) |  | 29.02 | (28.92,29.11) |  | 6.94 | (6.89,6.99) |  | 48.09 | (47.99,48.20) |  |
| 55 | 967,776 | 73.15 | (73.06,73.24) |  | 28.29 | (28.20,28.38) |  | 6.81 | (6.76,6.86) |  | 47.94 | (47.84,48.04) |  |
| 60 | 1,055,755 | 72.55 | (72.46,72.63) |  | 27.56 | (27.48,27.65) |  | 6.68 | (6.63,6.73) |  | 47.77 | (47.68,47.87) |  |
| 65 | 1,143,735 | 71.88 | (71.80,71.96) |  | 26.73 | (26.65,26.81) |  | 6.52 | (6.47,6.57) |  | 47.58 | (47.49,47.67) |  |
| 70 | 1,231,715 | 71.12 | (71.04,71.20) |  | 25.86 | (25.78,25.93) |  | 6.33 | (6.29,6.38) |  | 47.30 | (47.22,47.39) |  |
| 75 | 1,319,694 | 70.31 | (70.23,70.39) |  | 24.96 | (24.89,25.04) |  | 6.12 | (6.08,6.16) |  | 46.98 | (46.90,47.07) |  |
| 80 | 1,407,674 | 69.37 | (69.30,69.45) |  | 24.05 | (23.98,24.12) |  | 5.90 | (5.86,5.94) |  | 46.62 | (46.54,46.71) |  |
| 85 | 1,495,654 | 68.31 | (68.24,68.39) |  | 23.11 | (23.04,23.18) |  | 5.69 | (5.65,5.72) |  | 46.19 | (46.11,46.27) |  |
| 90 | 1,583,633 | 67.10 | (67.03,67.17) |  | 22.16 | (22.10,22.23) |  | 5.46 | (5.43,5.50) |  | 45.66 | (45.59,45.74) |  |
| 95 | 1,671,613 | 65.70 | (65.62,65.77) |  | 21.20 | (21.14,21.26) |  | 5.24 | (5.20,5.27) |  | 45.01 | (44.94,45.09) |  |

Note: In the heading, the table depicts the proportion of ELM for each disadvantaged sub-group in the test set. In the rows, the table depicts, for each disadvantaged sub-group, the proportion of actual ELM captured by our model and their confidence intervals.

**Figure 3:**
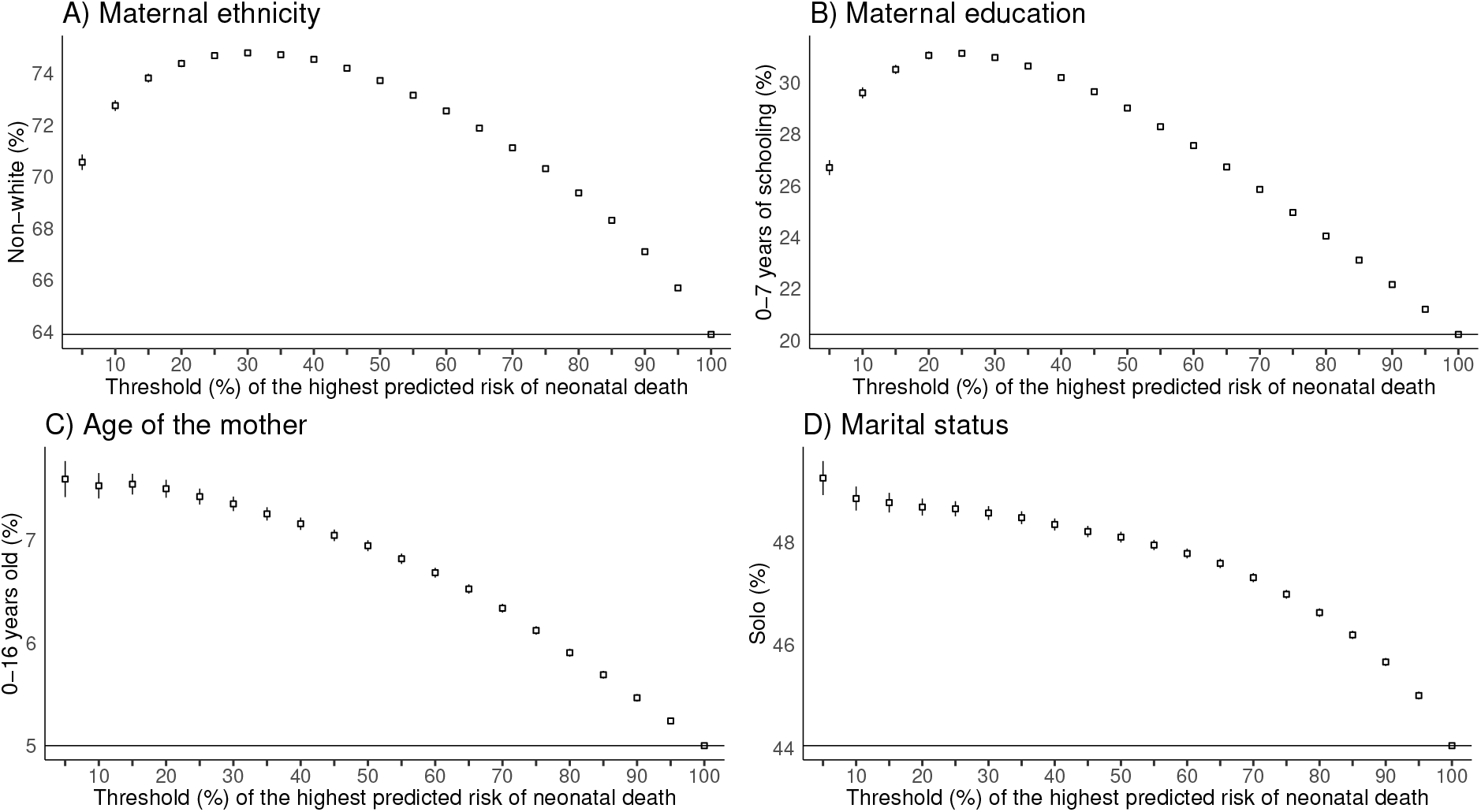
Proportion of individuals selected by the model. Note: In each graph, the horizontal lines depict the proportion of each ELM in the test set. The data points mark the proportion of actual ELM captured by our model and their confidence intervals.

The numbers in the table and the graphs in the figure demonstrate that our algorithm selects a significantly higher proportion of individuals from the disadvantaged sub-groups to be high risk for nearly all threshold percentages of the highest predicted risk. Only at the highest percentage thresholds, when the algorithm selects nearly the entire test set, does the proportion of disadvantaged sub-groups converge to the actual proportion in the test set. This means that the proportion of disadvantaged individuals selected by the algorithm is higher than the overall proportion in the test set.

One wonders whether the selection of these individuals would cause a distortion in the number of preventable deaths captured by the algorithm. Figure 4 shows the analysis of preventable deaths identified per subgroup. The analysis demonstrates that there are no statistical differences in the proportion of actual preventable deaths from disadvantaged populations that would be included in the selected at-risk births. Therefore, our preferred model is not biased against or in favor of underserved groups. On the contrary, using our algorithm would provide a fair inclusion for each population in terms of actual preventable deaths.

**Figure 4:**
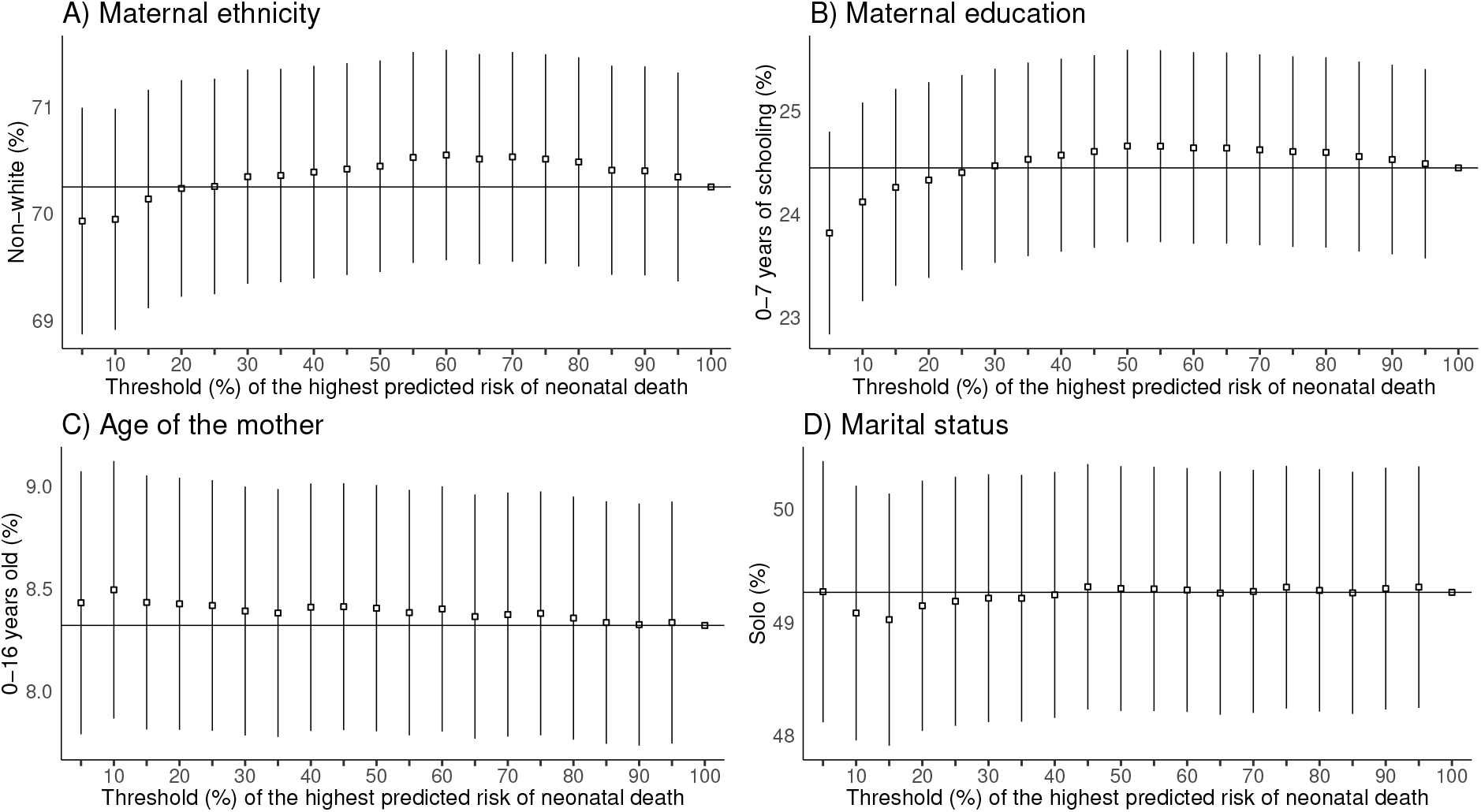
Proportion of actual ELM captured by the model. Note: In each graph, the horizontal lines depict the proportion of each ELM in the test set. The data points mark the proportion of actual ELM captured by our model and their confidence intervals.

**Figure 5:**
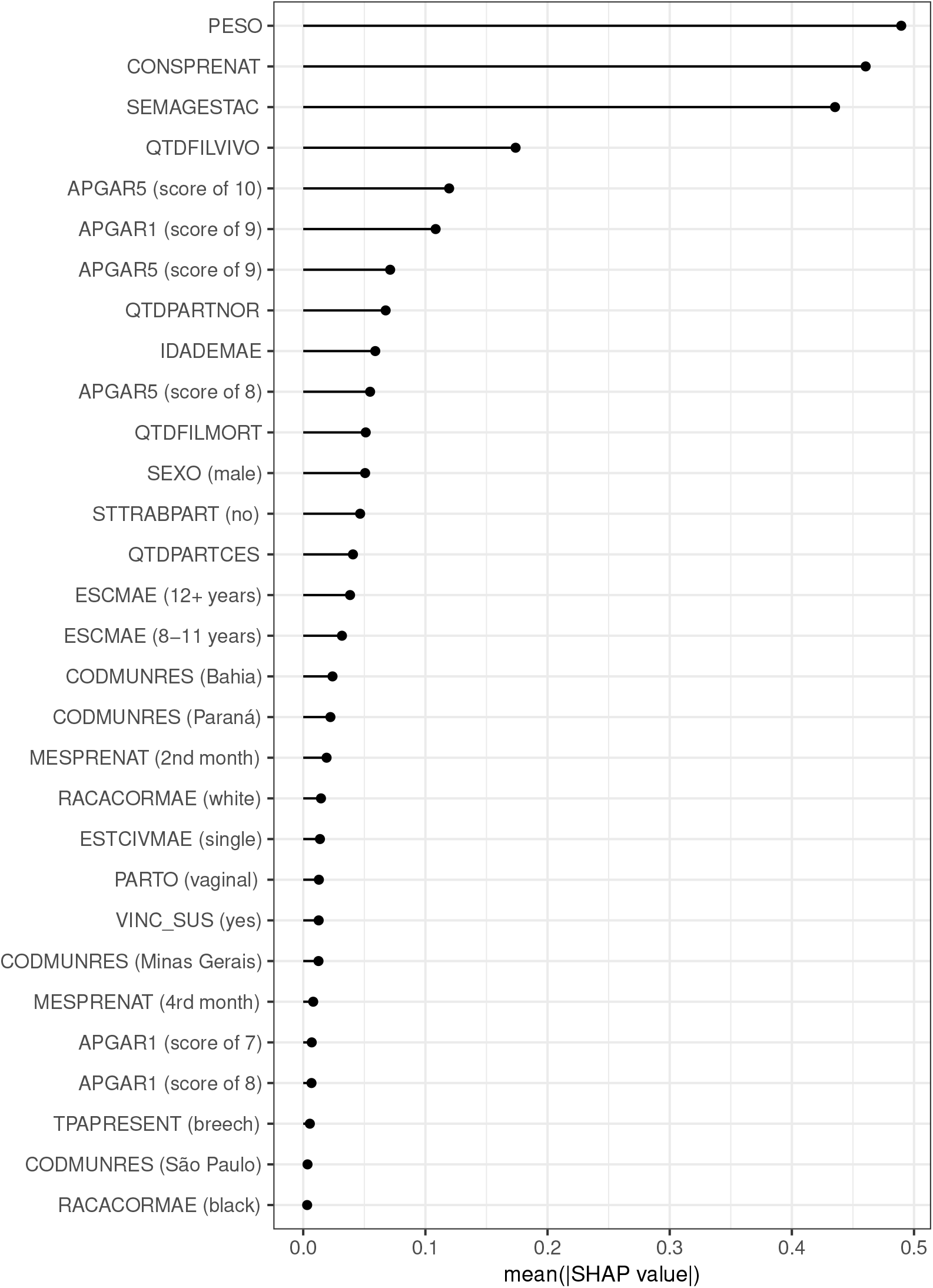
Feature importance based on Shapley Additive Explanations (SHAP) values. Note: The graph depicts the absolute mean SHAP values averaged over the various threshold percentages for the highest predicted risk of neonatal death. Variable names according to Table 1.

To emphasize the findings more clearly, Table 6 presents the outcomes of the hypothesis tests at a 95% confidence level. The null hypothesis is not rejected for all thresholds and sub-groups depicted in the table. It corroborates that using our algorithm would provide a fair inclusion for each population in terms of actual preventable deaths.

**Table 6:** Tests for proportions at a confidence level of 95%: comparing the proportion of preventable deaths captured by the algorithm and in the test sample.

| Threshold<br>(%) | $n$ | Maternal ethnicity<br>($p_0 = 70.25\%$) | | Maternal education<br>($p_0 = 24.45\%$) | | Age of the mother<br>($p_0 = 8.32\%$) | | Marital status<br>($p_0 = 49.26\%$) | |
| --- | --- | --- | --- | --- | --- | --- | --- | --- | --- |
| | | $\hat{p}$ (%) | p-value | $\hat{p}$ (%) | p-value | $\hat{p}$ (%) | p-value | $\hat{p}$ (%) | p-value |
| 5 | 7,221 | 69.93 | 0.5629 | 23.81 | 0.2173 | 8.43 | 0.7505 | 49.27 | 1.0000 |
| 10 | 7,591 | 69.95 | 0.5737 | 24.12 | 0.5126 | 8.49 | 0.7505 | 49.08 | 0.7591 |
| 15 | 7,753 | 70.14 | 0.8382 | 24.26 | 0.7089 | 8.43 | 0.7366 | 49.02 | 0.6798 |
| 20 | 7,866 | 70.23 | 0.9881 | 24.33 | 0.8183 | 8.42 | 0.7509 | 49.14 | 0.8419 |
| 25 | 7,945 | 70.25 | 1.0000 | 24.40 | 0.9361 | 8.42 | 0.7699 | 49.18 | 0.8971 |
| 30 | 8,030 | 70.34 | 0.8608 | 24.47 | 0.9762 | 8.39 | 0.8359 | 49.21 | 0.9352 |
| 35 | 8,087 | 70.35 | 0.8430 | 24.53 | 0.8723 | 8.38 | 0.8600 | 49.21 | 0.9340 |
| 40 | 8,143 | 70.39 | 0.7935 | 24.57 | 0.8061 | 8.41 | 0.7875 | 49.24 | 0.9768 |
| 45 | 8,188 | 70.42 | 0.7499 | 24.60 | 0.7475 | 8.41 | 0.7802 | 49.31 | 0.9379 |
| 50 | 8,219 | 70.44 | 0.7101 | 24.66 | 0.6638 | 8.40 | 0.7985 | 49.30 | 0.9604 |
| 55 | 8,252 | 70.52 | 0.5925 | 24.66 | 0.6657 | 8.38 | 0.8531 | 49.29 | 0.9650 |
| 60 | 8,282 | 70.55 | 0.5616 | 24.64 | 0.6912 | 8.40 | 0.8067 | 49.28 | 0.9787 |
| 65 | 8,319 | 70.51 | 0.6119 | 24.64 | 0.6928 | 8.36 | 0.9030 | 49.26 | 0.9996 |
| 70 | 8,345 | 70.53 | 0.5836 | 24.62 | 0.7189 | 8.37 | 0.8770 | 49.27 | 0.9970 |
| 75 | 8,363 | 70.51 | 0.6114 | 24.60 | 0.7460 | 8.38 | 0.8614 | 49.31 | 0.9423 |
| 80 | 8,386 | 70.48 | 0.6485 | 24.60 | 0.7584 | 8.35 | 0.9214 | 49.28 | 0.9830 |
| 85 | 8,408 | 70.40 | 0.7632 | 24.55 | 0.8247 | 8.33 | 0.9791 | 49.26 | 1.0000 |
| 90 | 8,430 | 70.40 | 0.7718 | 24.53 | 0.8722 | 8.32 | 1.0000 | 49.30 | 0.9601 |
| 95 | 8,444 | 70.34 | 0.8613 | 24.49 | 0.9410 | 8.33 | 0.9790 | 49.31 | 0.9410 |

The model underscores the importance of accounting for key individual risk factors—such as literacy, age, race, and marital status—to more effectively identify children at highest risk. With 85% of preventable neonatal deaths concentrated within the top 5% risk group, the algorithm demonstrates strong predictive power, reinforcing the value of machine learning in public health targeting.

Extensive subgroup analyses across ethnicity, education, age, and marital status reveal statistical parity in model outputs. This helps address concerns about algorithmic bias and supports ethical implementation. Crucially, fairness here refers not to targeting itself, but to the accurate identification of risk-outcome relationships, which is the core objective of our approach.

For example, to target the top 5% risk group, approximately 8% of underage mothers should be included—compared to their 5% representation in the overall dataset. Likewise, the model recommends including 50% of single mothers, even though they comprise only 44% of the population.

Finally, to exemplify the predictions, Table 7 provides examples of births classified as high, medium, and low-risk of preventable deaths. These examples illustrate the role of birth weight and Apgar scores as indicators of high risk, whereas we see that the lowest-risk births differentiate from the medium-risk births by demographics, such as years of schooling, and are associated with adequate birth weight and maternal ethnicity.

**Table 7:** Examples of birth profiles and risk predictions.

| Feature | High Risk |  | Medium Risk |  | Low Risk |  |
| --- | --- | --- | --- | --- | --- | --- |
| LOCNASC | Hospital | Other | Hospital | Hospital | Hospital | Hospital |
| VINC_SUS | Yes | Yes | Yes | Yes | Yes | No |
| IDADEMAE | 24 | 23 | 20 | 17 | 31 | 33 |
| SEXO | Female | Male | Male | Female | Female | Female |
| APGAR1 | 2 | 4 | 8 | 9 | 9 | 8 |
| APGAR5 | 3 | 8 | 9 | 10 | 10 | 10 |
| PESO | 1050 | 5250 | 3820 | 2980 | 3320 | 3855 |
| SEMAGESTAC | 44 | 25 | 39 | 40 | 38 | 40 |
| GRAVIDEZ | Single | Single | Single | Single | Single | Single |
| PARTO | Vaginal | Vaginal | Vaginal | Vaginal | Vaginal | Cesarean |
| ESCMAE | 8-11 years | 4-7 years | 1-3 years | 8-11 years | 12 or more | 12 or more |
| IDANOMAL | No | No | No | No | No | No |
| RACACORMAE | Brown | Brown | Brown | Brown | White | White |
| CONSPRENAT | 47 | 8 | 7 | 5 | 13 | 11 |
| MESPRENAT | 10 | 2 | 3 | 5 | 1 | 2 |
| TPAPRESENT | 1 | 1 | 1 | 1 | 1 | 1 |
| STTRABPART | 2 | 1 | 2 | 2 | 2 | 2 |
| TPNASCASSI | 1 | 1 | 1 | 1 | 1 | 1 |
| QTDFILVIVO | 0 | 0 | 2 | 0 | 2 | 10 |
| QTDFILMORT | 0 | 0 | 0 | 0 | 1 | [0] |
| QTDGESTANT | 1 | 1 | 3 | 1 | 4 | 2 |
| QTDPARTNOR | 1 | 1 | 3 | 1 | 17 | 1 |
| QTDPARTCES | 1 | 1 | 1 | 1 | 1 | 2 |
| ESTCIVMAE | Single | Civil union | Married | Single | Married | Married |
| CODMUNRES | BA | BA | CE | PE | RS | SP |

Note: Examples selected near the 1% highest risks (High Risk), 50% risks (Medium Risk), and 100% risks (Low Risk).

## 4 Discussion

Our findings demonstrate that policymakers and government agencies can harness existing data to facilitate more precise and cost-effective targeting of interventions. We accomplish this by developing a new analytic approach that integrates large administrative datasets and pairs them with ML models to enable the identification, with a high degree of precision, of births with the highest risk of preventable deaths. The level of accuracy of these models underscores their potential for application in health policy, particularly as early screening tools to identify neonates at elevated risk of preventable deaths. Using these models, policy-makers can develop proactive public health initiatives that aim to assess and reduce factors that contribute to infant mortality.

Our objective is to help local health professionals and policymakers identify which children need special attention, not based on preconceived risk factors, but by using a data-driven approach that combines several risk factors and provides digested information to healthcare providers or policymakers about those neonates who need more attention.

This is particularly useful, for example, in Brazilian regions where teams in the public health care system (SUS) can be responsible for 2000 to 3500 individuals^29^, as this identification might be very challenging due to the sheer number of patients under their care. The SUS is the world’s largest government-run public healthcare system by number of beneficiaries, land area coverage, and affiliated network with more than one million healthcare providers^30^. Based on our methodology, the use of an easy-to-use app ^2^ could assist healthcare teams on the ground in their targeting strategies by assigning a risk score for each neonate under their care.

To better accomplish our goal, we apply a new metric to evaluate the performance of the machine learning models developed, which is appropriate for public health professionals and policymakers. Many ML algorithms’ performance is judged by criteria such as specificity and accuracy, or F1 metrics that are difficult to interpret for policy purposes. Our metric evaluates the usefulness of a given ML algorithm to identify high-risk births from preventable causes for any given targeting coverage threshold.

The rationale for our metric is the principle that life-saving interventions are only effective when directed toward individuals who would otherwise face fatal outcomes—making accurate identification of at-risk cases essential. Even a “miracle drug” that can counteract any cause of death can only reduce mortality if given to children who, without it, would have died. Because of this, interventions that cannot be given universally must be carefully targeted to those at the highest risk of mortality (absent the intervention) to have an efficient effect. Our method addresses this issue by ranking births by their risk of preventable deaths.

One of the main concerns of applying these types of machine-learning models is their potential to exacerbate existing socioeconomic inequalities ^31^. This can occur when predictive models are trained in ways that consistently lead to poorer performance for marginalized populations^32^. Our model does not suffer from this concern, as it properly selects a statistically equal proportion of births with preventable deaths for disadvantaged and privileged populations, muffling concerns of algorithmic bias. Bias here refers not to the targeting itself, but to the accurate identification of risk-outcome relationships, which is the core objective of our approach.

More specifically, our approach avoids bias in favor of more privileged groups by selecting a higher proportion of disadvantaged individuals—relative to ELM population averages—to achieve equitable outcomes in terms of preventable deaths. Policymakers can use the detailed results we produced (in the aforementioned Table 5) to guide an equitable allocation of health interventions. For instance, to target the top 5% risk group in an equitable way, approximately 71% of non-white mothers should be included compared to their 64% representation in the overall dataset. Likewise, the model recommends including 49% of single mothers, even though they comprise only 44% of the population.

We emphasize that any ML application in healthcare should undergo a thorough evaluation and discussion with domain experts before implementation. Our approach is not a replacement for health care professionals, who have subject matter expertise that should not be ignored. Instead, we are offering one additional tool for them. This tool can be particularly useful in situations, such as those in which healthcare teams are responsible for a large number of patients.

As a caveat, our study presumes that healthcare providers can make the right intervention to save at-risk newborns. That is the reason behind ranking the risk of death from preventable causes. Of course, for this to become true, it depends on the actual capacity of the policymaker or healthcare provider to intervene correctly in preventable death cases.

Our approach aligns with recent trends in clinical medicine and health policy on the use of predictive modeling to aid and better inform decision-making. ^33,34,35,36,37^. It is also in line with recent trends in personalized medicine, which is gaining prominence in other fields of medicine and public health, as risk assignments are estimated at the individual level^38,39,40^.

Other papers focus on early-life mortality prediction in Brazil^41,42^ and in other Low and Middle Income countries, such as Iran ^43^ or sub-Saharan Africa ^44^. We add to these contributions by focusing on mortality from preventable causes, adopting a performance metric that is more useful for policy and clinical decision making, and by developing and testing an approach that does not discriminate against disadvantaged populations.

## 5 Conclusions

Using publicly accessible administrative data and machine learning techniques allows for a highly accurate identification of births at the highest risk of preventable mortality. This is useful for policymakers as they can target health interventions to those who need the most and where they can be effective without producing bias against disadvantaged populations. Taken together, our findings show that a risk-scoring algorithm needs to select a higher proportion of individuals from the less-advantaged populations as high-risk births to provide a statistically equal proportion of births with preventable deaths for both disadvantaged and privileged sub-groups.

Our approach can guide policymakers in reducing neonatal mortality rates and help them intervene in preventable death cases correctly and unbiasedly. The methods and metrics developed in this paper have wide applicability and are flexible enough to apply to several scenarios in other developing countries. For example, some countries with incomplete vital registration systems could use surveys like the Health and Demographic Surveys (DHS) instead of administrative data. Inclusion of risk factors can also vary between countries, given the availability of data and political and public health considerations.

## 6 Declarations

### 6.1 Ethics approval and consent to participate

The study was approved by the Getulio Vargas Foundation (FGV) Committee for Ethics in Research Involving Human Subjects.

### 6.2 Consent for publication

None declared.

### 6.3 Data availability

The datasets analysed during the current study are available in the DATASUS repository, https://datasus.saude.gov.br/.

### 6.6 Authors’ contributions

Raphael Saldanha performed the feature engineering; Marcus Nascimento performed the main statistical analyses; Antonio Ramos and Fabio Caldieraro planned the research, designed the study, and wrote the main manuscript.

## 6.7 Acknowledgements

The authors thank Chad Hazlett, PhD, and participants of the EBAPE Graduate Seminars and FJLES Research Seminars for comments on an early version of this paper, and Isabella Grion for her help in editing the paper.

## Footnotes

1 We note that we also performed linkages using probabilistic matching using the method proposed by ^15^. Although the probabilistic matching enabled us to link a larger proportion of records, we found later in the analysis that the use of probabilistic matching did not improve the predictive power of our algorithms; thus, for the sake of parsimony, we used the dataset with deterministic matching. Nevertheless, we comment that analyses performed in other settings may benefit more from probabilistic matching than our analysis did.

2 We developed an example of such an app and made it available on the Internet at https://64o4b7-marcus0l0nascimento.shinyapps.io/tent_app/

## References

1 Johnson RC, Schoeni RF. The Influence of Early-Life Events on Human Capital, Health Status, and Labor Market Outcomes Over the Life Course. The BE Journal of Economic Analysis & Policy. 2011;11(3).

2 Currie J, Vogl T. Early-Life Health and Adult Circumstance in Developing Countries. Annual Review of Economics. 2013;5:1–36.

3 Smith LK, Manktelow BN, Draper ES, Springett A, Field DJ. Nature of socioeconomic inequalities in neonatal mortality: population based study. BMJ. 2010;341:c6654.

4 Dyer L, Theall KP, Wallace M. Structural racism, racial inequities and urban–rural differences in infant mortality in the US. Journal of Epidemiology & Community Health. 2020;75(8):788–793.

5 Bhutta ZA, Das JK, Bahl R, Lawn JE, Salam RA, Paul VK, et al. Can available interventions end preventable deaths in mothers, newborn babies, and stillbirths, and at what cost? Lancet. 2014;384(9940):347–370.

6 Deep M. E-Economic and Social Council-Report of the Inter-Agency and Expert Group on. 2016;.

7 Chao F, You D, Pedersen J, Hug L, Alkema L. National and regional under-5 mortality rate by economic status for low-income and middle-income countries: a systematic assessment. The Lancet Global Health. 2018;6(5):e535–e547.

8 Ramos AP, Weiss RE. Measuring Within and Between Group Inequality in Early-Life Mortality Over Time: A Bayesian Approach with Application to India. arXiv preprint 180408570. 2019;.

9 Ramos AP, Flores MJ, Weiss RE. Leave no child behind: Using data from 1.7 million children from 67 developing countries to measure inequality within and between groups of births and to identify left behind populations. PLOS ONE. 2020;15(10):e0238847.

10 Panch T, Mattie H, Atun R. Artificial intelligence and algorithmic bias: implications for health systems. Journal of Global Health. 2019;9(2):020318.

11 Obermeyer Z, Vogeli BPC, Mullainathan S. Dissecting racial bias in an algorithm used to manage the health of populations. Science. 2019;60(6464):447–453.

12 Kakani P, Chandra A, Mullainathan S, Obermeyer Z. Allocation of COVID-19 Relief Funding to Disproportionately Black Counties. JAMA. 2020;324(10):1000–1003.

13 Akter S, McCarthy G, Sajib S, Michael K, Dwivedi YK, D’Ambra J, et al. Algorithmic bias in data-driven innovation in the age of AI. International Journal of Information Management. 2021;60:102387.

14 Mullainathan S, Obermeyer Z. Diagnosing Physician Error: A Machine Learning Approach to Low-Value Health Care. Quarterly Journal of Economics. 2022;137(2):679–727.

15 Enamorado T, Fifield B, Imai K. Using a probabilistic model to assist merging of largescale administrative records. American Political Science Review. 2019;113(2):353–371.

16 Batista AFM, Diniz CSG, Bonilha EA, Kawachi I, Filho ADPC. Neonatal mortality prediction with routinely collected data: a machine learning approach. BMC Pediatrics. 2021;21(322).

17 Malta DC, Duarte EC, de Almeida MF, de Salles Dias MA, de Morais Neto OL, de Moura L, et al. Lista de causas de mortes evitáveis por intervenções do Sistema único de Saúde do Brasil. Epidemiologia e Serviços de Saúde. 2007;16(4):233–244.

18 de Freitas Saldanha R, Bastos RR, Barcellos C. Microdatasus: pacote para download e pré-processamento de microdados do Departamento de Informática do SUS (DATASUS). Cadernos de Saúde Pública. 2019;35(9):1–9.

19 Honaker J, King G, Blackwell M. Amelia II: A Program for Missing Data. Journal of Statistical Software. 2011;45(7):1–47.

20 R Core Team. R: A Language and Environment for Statistical Computing. Vienna, Austria; 2021. Available from: https://www.R-project.org/.

21 Kleinbaum DG, Klein M. Logistic Regression. 1st ed. New York: Springer; 2010.

22 Tibshirani R. Regression Shrinkage and Selection Via the Lasso. Journal of the Royal Statistical Society, Series B. 1996;58(1):267–288.

23 Zou H, Hastie T. Regularization and variable selection via the Elastic Net. Journal of the Royal Statistical Society, Series B. 2005;67:301–320.

24 Breiman L. Random Forests. Machine Learning. 2001;45:5–32.

25 Chen T, Guestrin C. XGBoost: A Scalable Tree Boosting System. In: Proceedings of the 22nd ACM SIGKDD International Conference on Knowledge Discovery and Data Mining. KDD ‘16. New York, NY, USA: ACM; 2016. p. 785–794. Available from: http://doi.acm.org/10.1145/2939672.2939785.

26 Breiman L, Friedman JH, Olshen RA, Stone CJ. Classification and Regression Trees. Monterey, CA: Wadsworth and Brooks; 1984.

27 Aggarwal CC. Neural Networks and Deep Learning: A Textbook. 1st ed. Cham: Springer; 2018.

28 Lundberg SM, Lee SI. A unified approach to interpreting model predictions. Advances in neural information processing systems. 2017;30.

29 Ministério da Saúde. Portaria nº 2.436; 2017-09-21. Diário Oficial da União.

30 Castro MC, Massuda A, Almeida G, Menezes-Filho NA, Andrade MV, de Souza Noronha KVM, et al. Brazil’s unified health system: the first 30 years and prospects for the future. The Lancet. 2019;395(10195):P345–356.

31 d’Elia A, Gabbay M, Rodgers S, Kierans C, Jones E, Durrani I, et al. Artificial intelligence and health inequities in primary care: a systematic scoping review and framework. Family Medicine and Community Health. 2022;10(Suppl 1):e001670.

32 Mhasawade V, Zhao Y, Chunara R. Machine learning and algorithmic fairness in public and population health. Nature Machine Intelligence. 2021;3(8):659–666.

33 Sackett DL, Rosenberg WMC, Gray JAM, Haynes RB, Richardson WS. Evidence based medicine: what it is and what it isn’t. BMJ. 1996;312(7023):71–72.

34 Sackett DL. Evidence-based medicine. Seminars in perinatology. 1997;21(1):3–5.

35 Giacomini M. Theory-Based Medicine and the Role of Evidence: Why the Emperor Needs New Clothes, Again. Perspectives in Biology and Medicine. 2009;52(2):234–251.

36 Bluhm R, Borgerson K. Evidence-Based Medicine. In: Gifford F, editor. Philosophy of Medicine. Elsevier; 2011. p. 203–238.

37 Djulbegovic B, Guyatt GH. Progress in evidence-based medicine: a quarter century on. The Lancet. 2017;390(10092):415–423.

38 Hamburg MA, Collins FS. The Path to Personalized Medicine. New England Journal of Medicine. 2010;363:301–304.

39 Hayes DF, Markus HS, Leslie RD, Topol EJ. Personalized medicine: risk prediction, targeted therapies and mobile health technology. BMC Medicine. 2014;125(37).

40 Hoeyer K. Data as promise: Reconfiguring Danish public health through personalized medicine. Social Studies of Science. 2019;49(4):531–555.

41 Beluzo CE, Silva E, Alves LC, Rodrigo Campos Bresan NMA, Sovat R, Carvalho T. Towards neonatal mortality risk classification: A data-driven approach using neonatal, maternal, and social factors. Informatics in Medicine Unlocked. 2020;20(10):e100398.

42 Moreira JR, Bernardino HS, Vieira AB. Predição de Ó bito Neonatal usando Dados dos Sistemas de Informação do SUS e de Censo Demográfico. In: Simpósio Brasileiro de Computação Aplicada à Saúde (SBCAS). SBC; 2022. p. 234–245.

43 Sheikhtaheri A, Zarkesh MR, Moradi R, Kermani F. Prediction of neonatal deaths in NICUs: development and validation of machine learning models. BMC Medical Informatics and Decision Making. 2021;21(131).

44 Ramos AP, Hazlett C, Smith S. Better individual-level risk models can improve the targeting and life-saving potential of early-mortality interventions. Scientific Reports. 2023;13(1):21706.

